# Study of the effectiveness of partial quarantines applied to control the spread of the Covid-19 virus

**DOI:** 10.1101/2021.04.03.21254727

**Authors:** A. León

## Abstract

In Chile and in many countries of the world, partial quarantines have been used as part of the strategy to contain and control the Covid-19 virus. However, there is no certainty of its effectiveness and efficiency due to the lack of comparison with similar scenarios. In this work, we formulated a theoretical model of individual mobility, which also incorporates the infection dynamics of Covid-19. The model is based on a cellular automaton, which includes individuals moving through the represented spatial region and interacting according to the dynamics of Covid-19. In addition, we include mobile and partial health barriers, and different mobility regimes. Our results show that, partial quarantines would not be effective in general, to reduce the peak of active individuals infected with the virus, except for some proportions of territorial area involved in the division of the global region. Another interesting result of our research is that the passage restrictions in a sanitary barrier would not be relevant to the impact of the pandemic indicators in a sanitary quarantine regime. A possible explanation for the ineffectiveness of partial quarantines lies in the fact that the sanitary barriers are permeable to infected individuals and therefore when one of these individuals passes, an outbreak occurs in the virus-free zone that is independent of the original one.

## 1. Introduction

Chile has been one of the countries most affected by the Covid-19 pandemic and is among the first 15 places in the total number of registered cases and among the first 20 places regarding the number of cases per million people [1, 2]. The health authorities, since the beginning of the pandemic in the country, have based the strategy to combat it on two fundamental pillars: massive tests and dynamic and partial quarantine [3]. Evaluating the impact of the dynamic and partial quarantine strategy, in real time, is extremely difficult, as we do not have a control group, or a similar situation elsewhere and at another time. When the pandemic is occurring, the health authorities can change the limits of the quarantined areas and the level of confinement, depending on the incidence of cases and some other variables related to the place and the quarantined population, For example, the availability of hospitals and medical centers [4]. However, there is no theoretical platform that allows us to have knowledge of the optimal combinations of level of confinement, level of permeability of sanitary cords, and the extent of these measures over time.

We are going to define a partial and dynamic quarantine, as follows: A region of space, could be a city, is divided into subregions, with clearly defined limits. These subregions will have different levels of mobility (partial quarantine) and the limits of these subregions will have different levels of permeability. The boundaries of the regions and the levels of confinement of these regions change over time (dynamic quarantines).

In this work, we formulate a model to simulate a partial and dynamic quarantine and to assess the impact, in different propagation scenarios of Covid-19. Our results show that in general, partial quarantines would not be effective to reduce the peak of active individuals infected with the virus, except for some proportions of territorial area involved in the division of the global region. Another interesting result of our research would be that the passage restrictions in a sanitary barrier would not be relevant in the impact of the indicators of the pandemic in a sanitary quarantine regime. A possible explanation for the ineffectiveness of partial quarantines lies in the fact that the sanitary barriers are permeable to infected individuals and therefore when one of these individuals passes, an outbreak occurs in the virus-free zone that is independent of the original one. These new outbreaks follow the usual dynamics of virus spread and therefore increase the infection indicators in the total area.

## 2. Model

Our model is based on the theoretical platform of cellular automata. Cellular automata are models used to simulate the dynamics of complex systems. The main idea is to define an array of entities (cells of the automaton) that have a discrete set of states. The change of state of a particular cell is defined with updating rules for the automaton and depends on the state of the considered and neighboring cells. A variety of models based on CA have been used to efficiently study problems in biology, physics, chemistry, engineering, and materials sciences [5, 6, 7, 8, 9]. They represent an excellent alternative to models based on differential equations and Monte Carlo algorithms because they can simulate highly complex systems with a low computational cost.

Our cellular automaton model will be applied to Chilean cities, and therefore we will briefly discuss Chile’s political division. In Chile, the political division of the national territory is as follows: the country is divided into regions, the regions into provinces and the provinces, into municipalities. A large city, like Santiago de Chile, has more than 30 municipalities, and therefore, we will take the municipality as the natural territorial division of our model.

The model formulated in this work considers a spatial region that represents one or more municipalities. This region is divided into elemental cells, which form the cellular automaton. Over this spatial region, *n* particles moving through the CA are considered. These particles represent individuals. This technique, which consists of mixing a cellular automaton with particles that move through it, is known as “Lattice-gas cellular automaton, (LGCA)” and has been used to study the dynamics of diverse populations in biology and in dynamic systems in general [10, 11, 12]. In our model, the velocity of the particles is incorporated, through velocity channels that each elemental cell has. In figure 2.2 a), a unit cell of the CA is shown, which has five speed channels. The direction of each channel is also indicated in the same figure and we consider a channel with zero speed. This cell represents, a space within each municipality, a house, a market, a building, a hospital, etc. This allows simulating the effect of quarantined population, that is, a number of particles at rest, in channel number five.

**Figure 2.1:**
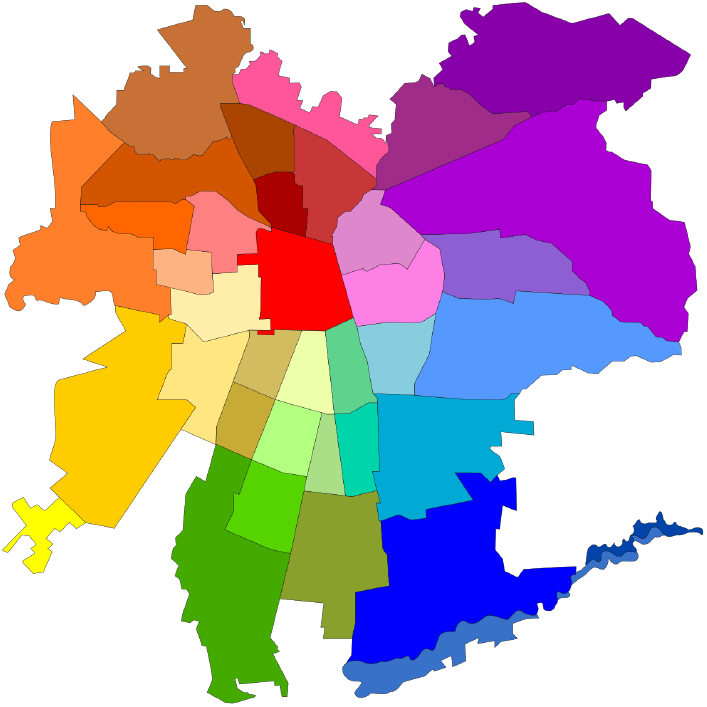
Municipalities represent the smallest region of political division in the city of Santiago de Chile.

**Figure 2.2:**
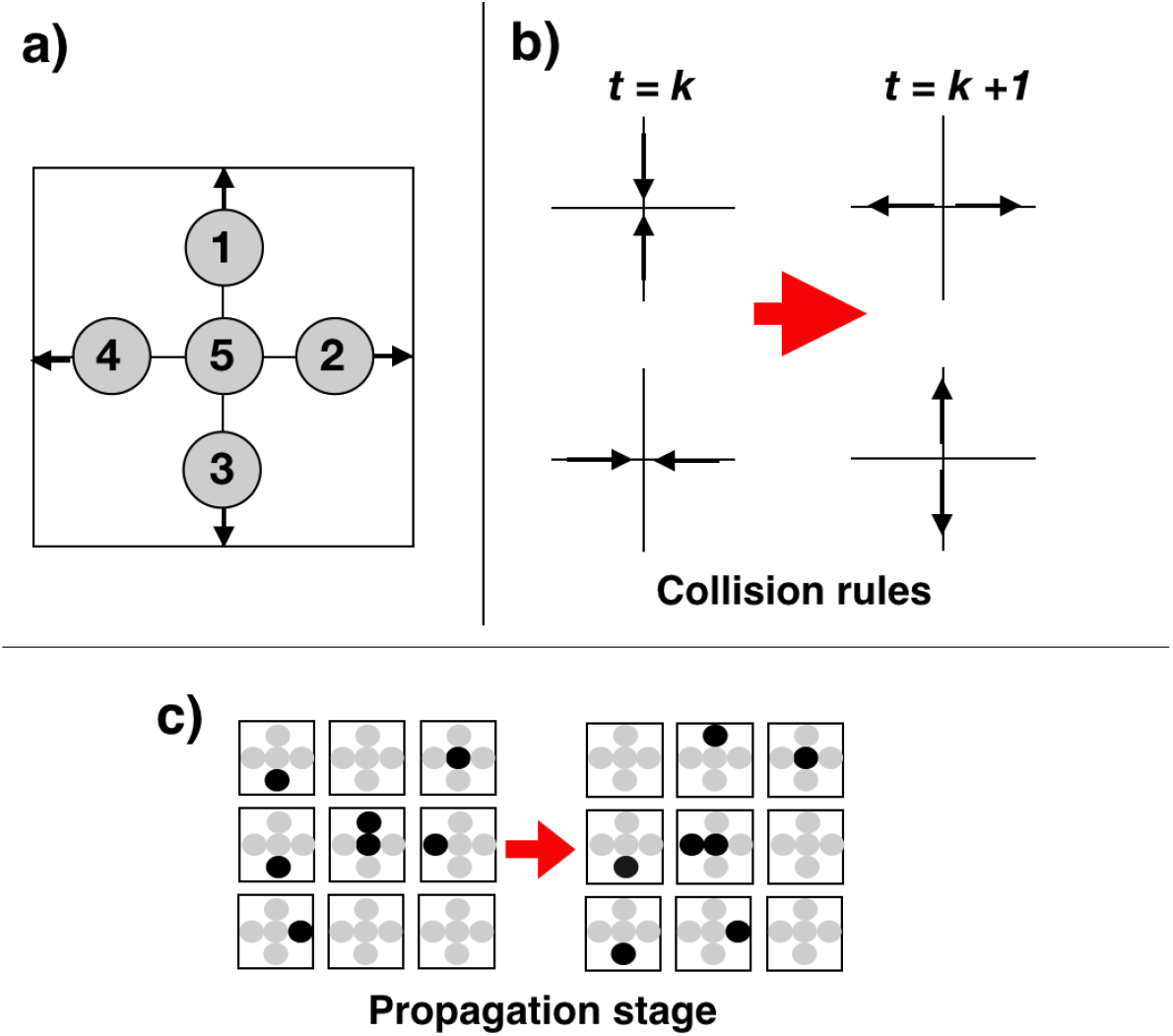
a) Diagram representing an elementary cell of the CA, with five speed channels. b) Collision rules, used in particle dynamics. c) Propagation stage. The cells on the left show the particle configuration at time *t*_1_ = *k*, while the cells on the right show the particle configuration at time *t*_2_ = *k* + 1.

The state of each cell, at time t, is specified through a five-component state vector *Ψ* = (*n*_1_, *n*_2_, *n*_3_, *n*_4_, *n*_5_). The components *n*_*i*_, with *i* = 1..5, of the state vector Ψ, correspond to the occupation number of each channel, in the cell considered. When the occupation number *n*_*i*_ is zero, then the corresponding speed channel is empty. If *n*_*i*_ is not zero, then it indicates that in the velocity channel, there is an individual. If this number is equal to one, *n*_*i*_ = 1, then the individual is susceptible to Covid-19 infection. If the occupation number is between 1 < *n*_*i*_ < *t*_*max*_, the individual is infected and is a contagious individual. Finally, if the occupation number *n*_*i*_ ≥ *t*_*max*_, the individual recovers and is therefore not infectious. The size of the timeslot and the occupation number in the state vector, are integers. We will return to this point, when we explain the dynamics of the CA.

### 2.1. Evolution of the cellular automaton

The dynamics of our CA contains two parts. The first one corresponds to the application of the Covid-19 infection operator and the second part corresponds to the movement of individuals (particles). In turn, the part that involves the movement of the particles is divided into a collision and a propagation stages. The collision phase is carried out between the particles themselves, when appropriate, and between particles and barriers, which represent the limits of the considered spatial region.

#### 2.1.1. Covid-19 infection operator

For each time step, the occupation number of the five channels of each of the cells of the CA is read. If any channel is occupied by a contagious individual, the cell is considered contagious and if there are susceptible individuals in the remaining channels, from the same cell, they are infected with probability *p*. The occupation number of the channels with individuals that became infected in this time step and of the channels with infectious individuals, increases by one, *n*_*i*_ = *n*_*i*_ + 1. When the occupation number of a contagious individual reaches the value *t*_*max*_, then he becomes a recovered individual. If there is a channel with a recovered individual in the inspected cell, then the infection operator is not applied to this channel.

#### 2.1.2. Movement of individuals

The movement of individuals, obeys a principle of exclusion, that is, a speed channel, can only contain one particle, or be empty. The first phase of the movement corresponds to the collision stage. In the LGCA models, which are used to study physical particle systems, there are collision operators, which must satisfy the linear momentum conservation principles. In our model, this condition is not necessary, however, we will maintain it, because it allows us to maintain a degree of randomness to the movement of individuals. These rules of collision between particles, which will be used in the present model, are represented in figure 2.2 b).

The propagation stage consists of the movement of the particles from the current cell to the neighboring cell, according to the corresponding speed. For example, suppose that a particle is in channel 1, of a given cell, at time *t*. This particle propagates to the neighboring cell, which is at the top of the current cell, occupying channel 1. In this propagation process, the exclusion principle is satisfied, because if in the destination cell, there were a particle, it will jump to the next cell, vacating the corresponding channel. Particles in channel 5 (zero speed) do not propagate. A schematic of this stage of propagation is shown in Figure 2.2 c). In our model, we have considered rigid barriers, at the limits of the spatial region under study. For these cells, this propagation rule is modified in such a way that the particles collide elastically, undergoing a change in their velocity of 180 degrees, if the exclusion principle allows it. If the particle cannot occupy the opposite speed channel, because it will be occupied by another particle, which comes behind, then the particle will jump to any other channel, from the neighboring cells, which will be vacant in the future time. We make this correction to satisfy the exclusion principle. Finally, sanitary barriers, separating two contiguous subregions, obey a rule similar to that of rigid barriers. The only difference is that there is a certain probability of crossing the barrier from one area to another. This probability depends on the level of restriction *b* of passage that is imposed on the sanitary barrier.

## 3. Results

### 3.1. Parameter space and proof of concept

We define a rectangular area of 10,000 cells, with an aspect ratio of 200 cells × 50 cells along the *x* and *y* axes as shown in Figure 3.1 a). We also define a sanitary barrier, which divides the area, as shown in the same figure and that can be located at any point on the *x*-axis.

**Figure 3.1:**
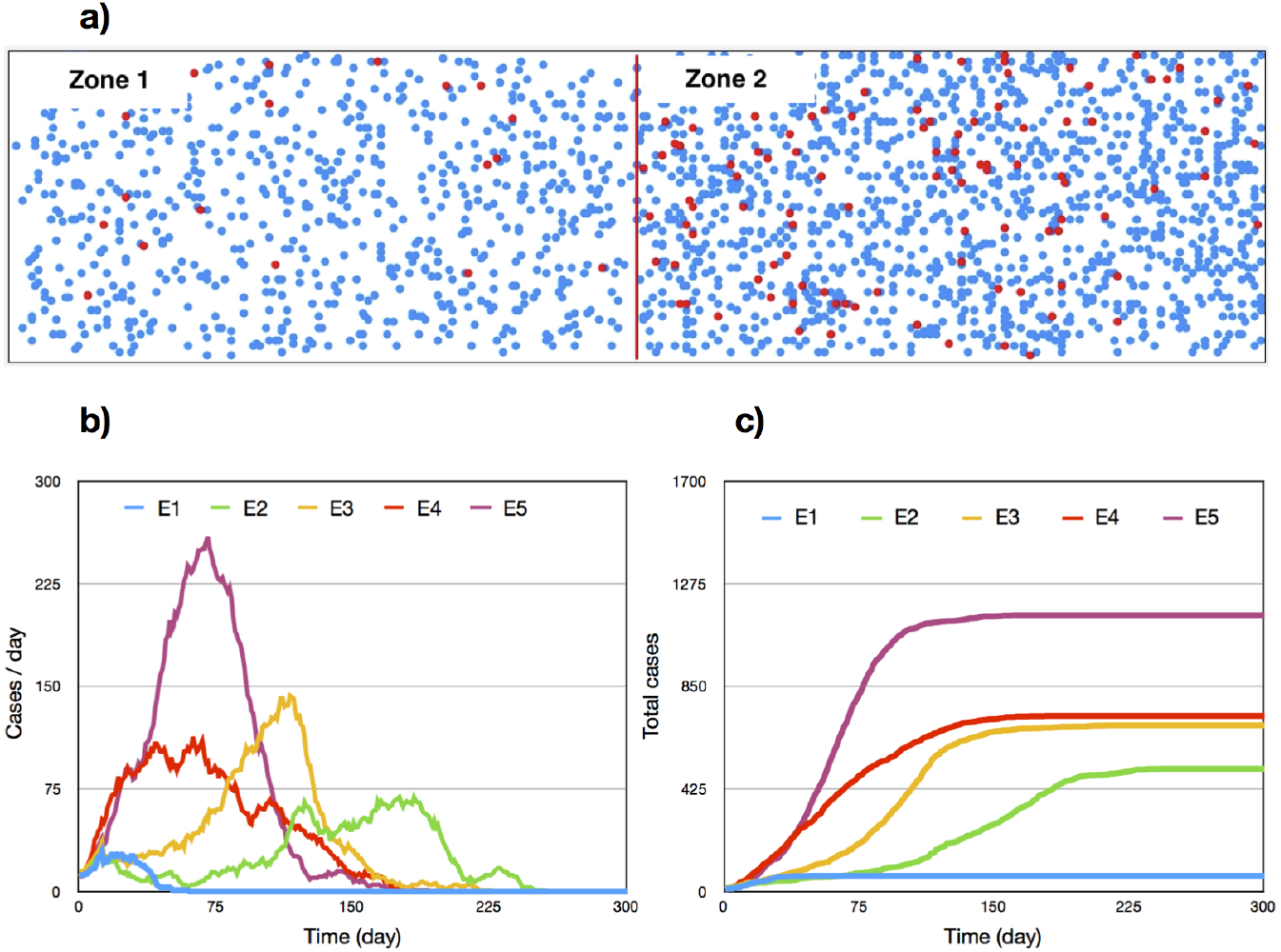
Simulation for five situations of the pandemic. In part a), the original rectangular region with 2,000 individuals is shown, separated by a sanitary barrier arranged right in the middle. Healthy or recovered individuals are shown in blue circles, while infected individuals are shown in red circles. Graphs b) and c) of this figure correspond to the infected cases per day and the total cases, considering the entire region.

Regarding the infection dynamics, in our model, consider close contact when an individual is with one or more individuals in the same cell. If there is an infected individual in that cell with the capacity to infect, the considered individual is infected with probability *p*. Furthermore, we define confinement (*c*) in our model, as the fraction of the population that does not move and, therefore, the individuals that remain in channel 5 of each cell. Remember that each channel can be empty or with only one individual. Let *c*_1_ and *c*_2_ be the confinements of zones 1 and 2, respectively. Finally, we define *b*, as the level of restriction in the sanitary barrier. This level is zero when all the individuals who need to cross the barrier can pass, and it is one when no individual passes. There are other additional parameters, such as the number of total individuals (*N*) in the simulation, the number of infected individuals (*N*_*i*_) at the start of the simulation and the position (*x*_*s*_) of the health customs. However, in this first phase of our study, these parameters will be fixed. The parameter space in the initial part of the study is shown in Table 1.

**Table 1:**
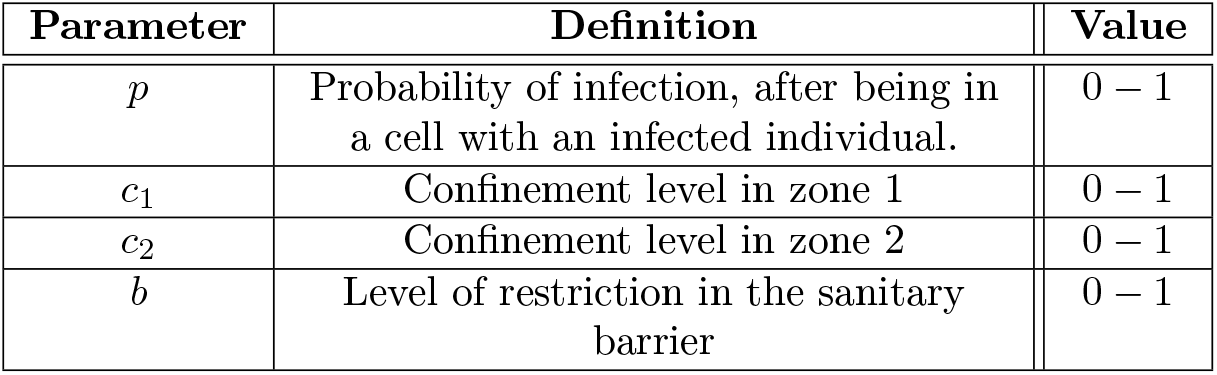
Parameter space in the first part of the study.

The first scenario that we are going to study corresponds to an area divided in half with the same number of individuals, (areas 1 and 2). The total number of individuals, considering both zones, is 2000. In addition, initially we considered 10 infected individuals randomly located in zone 2. We are going to consider a quarantine situation (almost total confinement) with *c* = 0.9, whereas a situation without quarantine, but with some movement restriction, it will be with *c* = 0.4. The justification of these extreme values for confinement *c* will be given in the next section of the results. Any value of the parameter *c*, intermediate between these two limits, will be an intermediate confinement.

Figure 3.1 shows the results of five simulations that will serve as a proof of concept, to guide our study in the parameter space. The initial conditions in the five cases studied for the fixed parameters are: *N* = 2, 000 *N*_*i*_ = 10 *x*_*s*_ = 100. The set of values of the four parameters of interest, for these five simulations is:

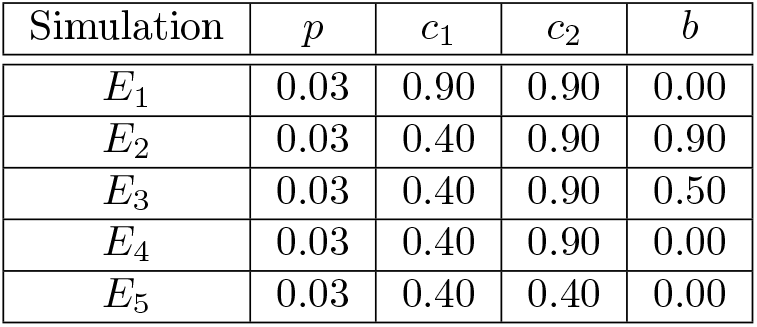

| Simulation | $p$ | $c_1$ | $c_2$ | $b$ |
| --- | --- | --- | --- | --- |
| $E_1$ | 0.03 | 0.90 | 0.90 | 0.00 |
| $E_2$ | 0.03 | 0.40 | 0.90 | 0.90 |
| $E_3$ | 0.03 | 0.40 | 0.90 | 0.50 |
| $E_4$ | 0.03 | 0.40 | 0.90 | 0.00 |
| $E_5$ | 0.03 | 0.40 | 0.40 | 0.00 |

In our results obtained with a single simulation, for each data set in the parameter space, we can observe two limit situations. These situations occur when we treat zones 1 and 2 as a single region without a sanitary barrier. Results E1 and E5, (blue and purple curves), show a situation of total quarantine (E1) and a situation of near normal mobility (E5). We can see that in situation E1, the outbreak can be controlled by reducing the mobility of the entire region, while in situation E5, the outbreak gets out of control, producing a wave of infections. However, the most important of this proof of concept corresponds to results E2, E3, and E4, which represent intermediate situations between a controlled outbreak and a severe wave of infections. In these three cases, zone 2 is quarantined and zone 1 is left with normal mobility. The difference between the three cases lies in the level of restriction in the sanitary barrier. As we can see in the three curves that describe the results E2, E3 and E4, as we increase the restriction of passage from one area to another, we lower the levels of contagion. This preliminary result raises the question of how efficient is it to manage the pandemic with dynamic and partial quarantines, applied in an area subdivided into smaller zones? In order to answer this question, we have developed a statistical study of the simulations and thus, sweep the parameter space in the search for this hypothetical optimal solution that reduces the level of contagion, without quarantining the entire region. We could find a combination of parameters that optimize the management of the pandemic, without having to quarantine the entire area.

## 4. Exploration of the parameter space and statistical validity of the results

### 4.1. Protocol in simulations

For each study of the set of values in the parameter space, the simulation is repeated thirty times and from this, we calculate the mean value of the maximum number of infections per day; the mean value of the time in which the maximum number of infections occurs; and the mean value of the total number of infected individuals. For each of these central descriptors, we calculate the standard error.

### 4.2. Indicators of infection, depending on the level of confinement of the entire area

Let us start with the response of the infection indicators, at the level of confinement of zones 1 and 2, considering these zones as a single territorial region. Figure 4.1 shows the number of infected per day, the accumulated number of infected and the time in which the maximum number of infected cases per day occurs. Bars representing the standard error of each measure are included in all curves. We can see that for graphs a) and b), the error bars are smaller than the data points. For the graph that represents the time in which the peak of cases occurs per day, (graph c), errors of greater magnitude can be seen, especially in the case of the probability of 0.02. The simulated system is the same as that presented in the proof of concept in section 3.1. In this case, for the whole study we have *b* = 0 and the level of confinement is the same for both zones. Consequently, zones 1 and 2 are considered as a single region. In this graph we see a study for four different cases of probability of infection. This parameter measures how contagious the virus strain considered is. The first thing we notice in graphs a) and b) of this figure is that for small confinement values, there is an increase in cases per day and in total cases. This behavior for small confinement values is explained by the fact that the contagion efficiency is not maximum if the mobility is close to its maximum value (*c* = 0). In the model, when the confinement is close to zero, very few individuals are in channel five, (individuals at rest). The latter decreases the probability that 2 or more particles are in the same cell at the same time. This behavior in which the infection levels increase, when the confinement increases, occurs up to a certain threshold value, (*c*_*t*_). From this threshold value, contagion levels decrease as confinement increases. Our results show that the threshold confinement, for all the probabilities of infection, is close to 0.25. This apparently unusual behavior of the model is present in the real situation of the virus. Suppose that very few or no individuals are at rest, the probability of a close encounter is less than when there is an appreciable number of individuals at rest. This would be representing the situation of the agglomerations for prolonged periods of time [**?**].

**Figure 4.1:**
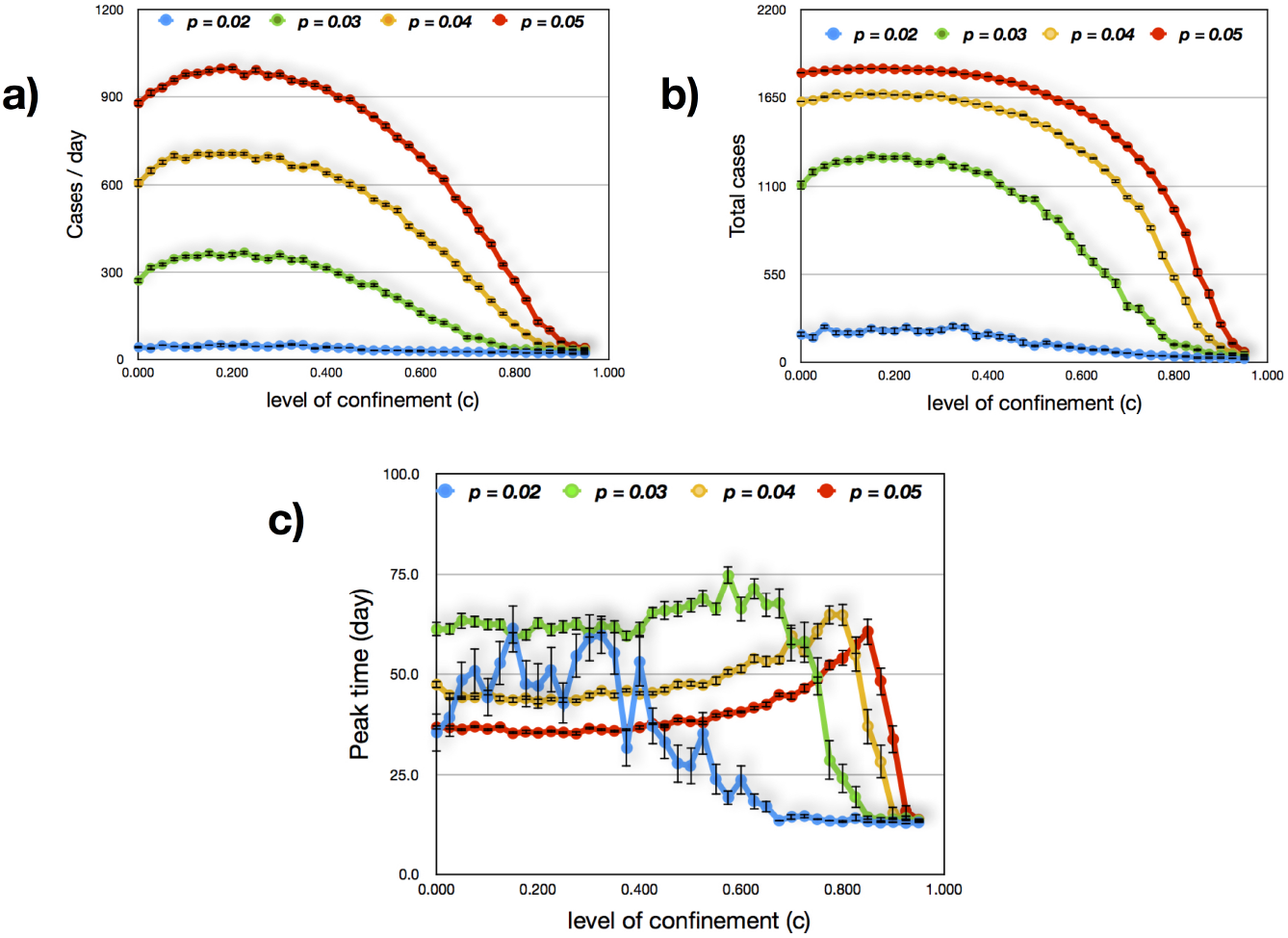
Study of infection indicators, depending on the level of confinement of zones 1 and 2, considered as a single region. a) Number of infections per day, b) Cumulative number of infections and c) Time in which the maximum number of infections per day occurs.

Considering that in times of the Covid-19 pandemic, classes in schools and universities were suspended in Chile and that many non-essential activities were stopped, even in cities and towns that were not in a pandemic, it is estimated that 40% [13, 14] of the population was in confinement and therefore, it is an amount that we will take as a confinement reference, for an area that is not in quarantine, *c* = 0.40.

According to estimates from the Chilean Ministry of Transportation, the traffic flow decreased, during the first wave of the epidemic, between 40% and 60% in the quarantined areas [13]. On the other hand, in a study on mobility that was carried out recently, with the data of customers of the mobile phone systems, it was established that during the quarantines, mobility decreased by values close to 40% in the quarantined towns [14]. Based on these estimates of pandemic mobility, the confinement limit in quarantine will be *c* = 0.90, in our model. Of course, our study contemplates confinement values, outside these limits, but it was necessary for our study to define theoretical mobility with and without quarantine. We can see in figure 4.1 that the number of cases per day and the total number of cases, for the confinement of *c* = 0.40, is close to the value with respect to c = 0 and this leaves us even more satisfied with the choice of this reference. At the other extreme, we can see that for *c* = 0.90, the outbreak is practically controlled for all infection probabilities.

Regarding the time in which the peak of cases per day occurs (graph c), it remains practically constant up to a high value of confinement, in which it decreases drastically. This decrease is due to the fact that for high confinement, the outbreak is controlled in the first phase of propagation.

### 4.3. Effect of partial quarantine on Covid-19 indicators

We have carried out a study, dividing the region into two zones. Zone 1 will be the region on the left and Zone 2 will be the region on the right. In all simulated cases, there is an initial outbreak of 10 infected individuals randomly located in zone 2. We will put the sanitary barrier in three different positions. These positions divide the total area in the following proportions: zone 1 will represent in this division: ¼, ½, and ¾ of the total area. In all the results that we show in this section, zone 1 will not be in quarantine (*c*_1_ = 0.40), while zone 2 will be in quarantine (*c*_2_ = 0.90). Under these conditions we simulate the development of the pandemic for different values of the sanitary barrier. In the three studies we present the number of cases per day, the total number of infected and the time of the peak in the cases per day. Furthermore, the standard error of each simulation is incorporated in all studies. The detail of the study of the covid-19 indicators, considered in this work, depending on the magnitude of the barrier, is discussed in Appendix A. As a summary of these results, we can say that the three indicators remain constant, regardless of the value of the sanitary barrier, except for values close to one, where the passage restriction is maximum. These changes start at values close to *b* = 0.8 and therefore we will take it as a reference of strict restriction of passage through the barrier.

Figure 4.2 shows the percentage reduction in the maximum value of cases per day and therefore the reduction in the peak of the outbreak, for a partial quarantine with the sanitary barrier located at x = 50. In this way, zone 2, where the outbreak is located, represents 75% of the total area considered. We incorporate the total quarantine reduction data into the figure, when zones 1 and 2 are considered as a single region, to help in comparing the effectiveness of partial quarantines. We can see in the figure that for the low probabilities of 0.2 and 0.3, the reduction in the peak is close to that reached when the quarantine is total. However, for the 0.4 and 0.5 probabilities it is less than the total quarantine reduction, but still significant. On the other hand, when the proportion on the surface of zone 2 is 50% and 25%, graphs 4.3 and 4.4 respectively, the reduction in the peak of infections per day is much less than the total quarantine regime. This reduction is negligible in the case of the barrier located at x = 150, even for a high value of the sanitary barrier.

**Figure 4.2:**
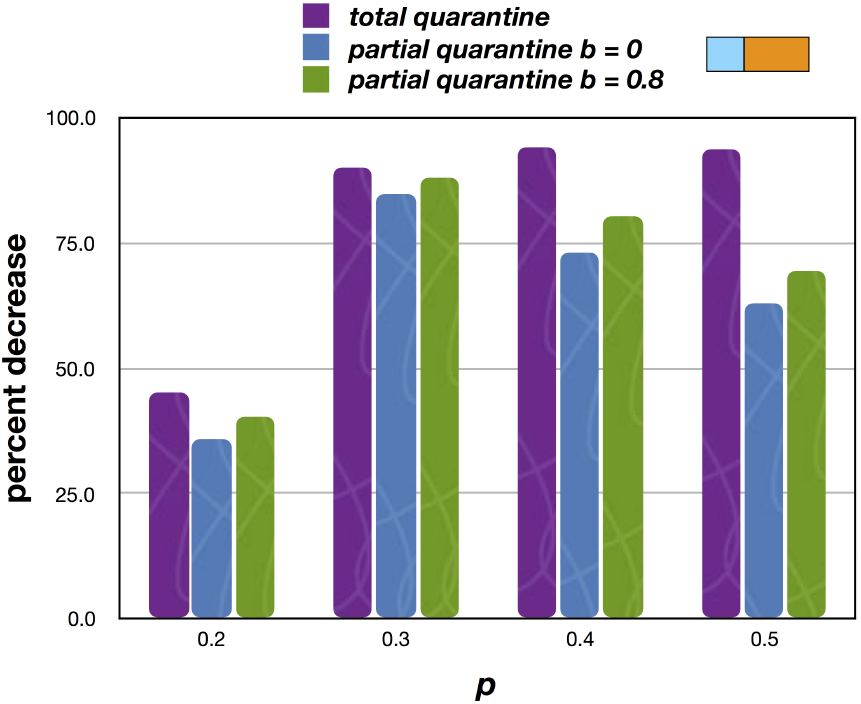
Percentage of reduction of the maximum value of cases per day, with respect to the epidemic without any type of quarantine, for the four contagion probabilities, considered in this study. The sanitary barrier is located at *x* = 50.

**Figure 4.3:**
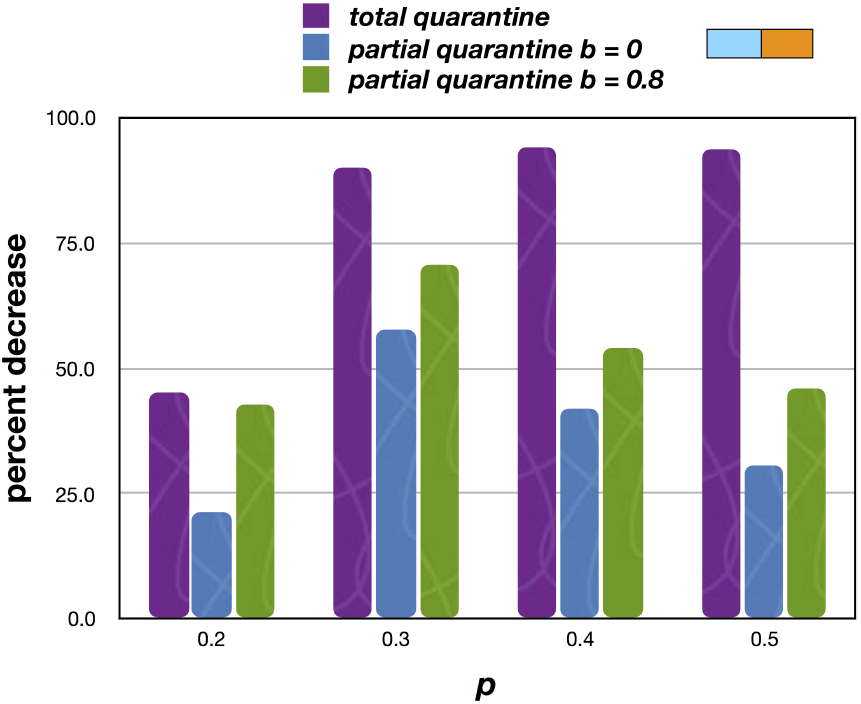
Percentage of reduction of the maximum value of cases per day, with respect to the epidemic without any type of quarantine, for the four contagion probabilities, considered in this study. The sanitary barrier is located at *x* = 100.

**Figure 4.4:**
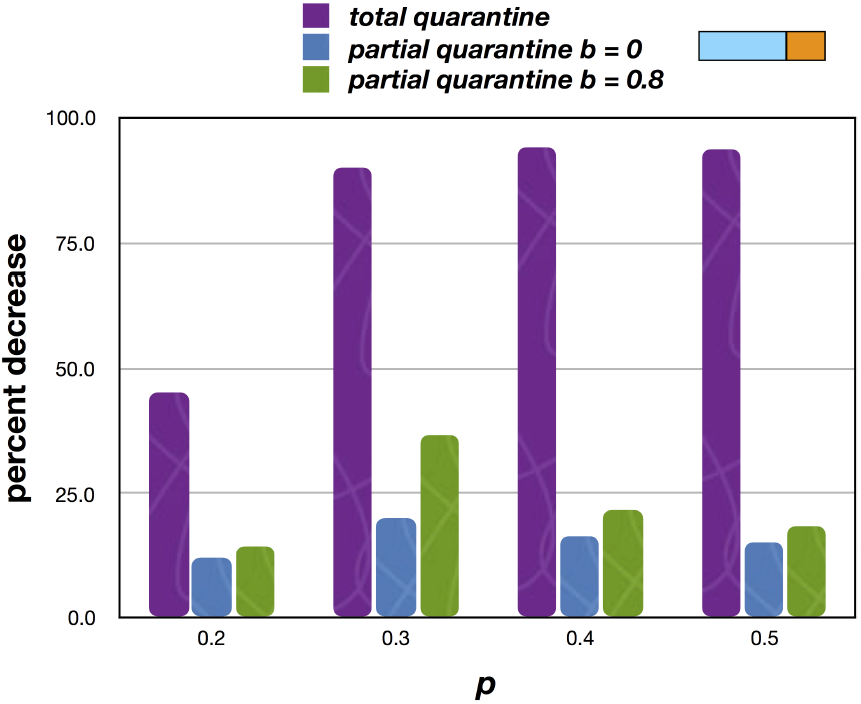
Percentage of reduction of the maximum value of cases per day, with respect to the epidemic without any type of quarantine, for the four contagion probabilities, considered in this study. The sanitary barrier is located at *x* = 150.

## 5. Conclusion

We have formulated a model based on cellular automata to study the spread of the Covid-19 virus, in different partial quarantine scenarios. In the results of the initial part of our proofs of concept, we have established that there could be some combinations of parameters for partial quarantines, which can control the epidemic. In the initial proof of concept it was established that when a region is divided into two zones, one where the Covid-19 outbreak begins and the other free of positive cases, imposing quarantine only on the area with the outbreak, in some cases manages to partially control the development of the epidemic. These preliminary results motivated us to deepen the study and scan the parameter space and look for some combination of these parameters with promising results in the control of the pandemic. In the second part of our research, we incorporated the systematic repetition of the simulations and we worked with the statistical descriptors of the mean value and the standard error. The mobility indicator was established in our model (*c*_1_ *and c*_2_), which would account for the reality in Chilean cities, regarding the quarantine regime and the normal mobility regime, during the first wave of infections. The results of this part of our study show that partial quarantines have very little effectiveness in controlling the development of the pandemic. It was established that when the regions considered have similar surfaces or when the region where the initial outbreak occurs has a smaller surface than the other region free of cases, partial quarantine is ineffective to control the pandemic, even for high values of the sanitary barrier. These results are valid for all the probabilities of infection evaluated in our study. In the particular case that the region with the initial outbreak has a larger surface area than the other case-free region, a partial and limited control of the pandemic is achieved. In our study, this case was registered with an area of the region with the outbreak of 75% of the total region. A possible explanation for the ineffectiveness of partial quarantines lies in the fact that the sanitary barriers are permeable to infected individuals and therefore when one of these individuals passes, an outbreak occurs in the virus-free zone that is independent of the original one. These new outbreaks follow the usual dynamics of virus spread and therefore increase the infection indicators in the total area.

## Data Availability

All data included in the article is publicly available

## Acknowledgments

To my son Alejandro León for his valuable contributions and his questions that served to enhance this research. To the Diego Portales University for its support to develop research in times of pandemic.

## 6. Appendix A

In this annex we show the results of the impact on the Covid-19 indicators that we have considered in our model, based on the health barrier that separates 2 regions of interest. The system studied is the same as that considered in section 4, that is, a region with 20,000 individuals divided into two regions. In one of them, (zone 2), there is an initial outbreak with 10 individuals randomly located in zone 2. This zone is quarantined (*c*_2_ = 0.9) and zone 1 is left with normal mobility (*c*_1_ = 0.4). The development of the outbreak is simulated, for different values of the sanitary barrier and for different positions of this barrier.

Figure 6.1 shows the results of the impact that the value of the sanitary barrier would have on the indicators of interest in our model when the barrier is located at x = 50. We can see that the three indicators considered do not change substantially with respect to the value of the health barrier and a significant change is only seen for values close to 1 in the health barrier. Values close to 1 are not realistic, since it would suppose isolation of the region where the outbreak occurs. In practice, the Chilean health authorities have based their partial quarantine strategy on the effectiveness of the sanitary barriers, called “sanitary cords”. However, we can see in the results provided by our model, the health barrier would have very little impact on the Covid-19 indicators, except for very high values of restriction of passage. This behavior is more acute when the barrier is located at *x* = 100 and *x* = 150, figures 6.2 and 6.3 respectively. With these proportions of surface, the impact of the sanitary barrier is less.

**Figure 6.1:**
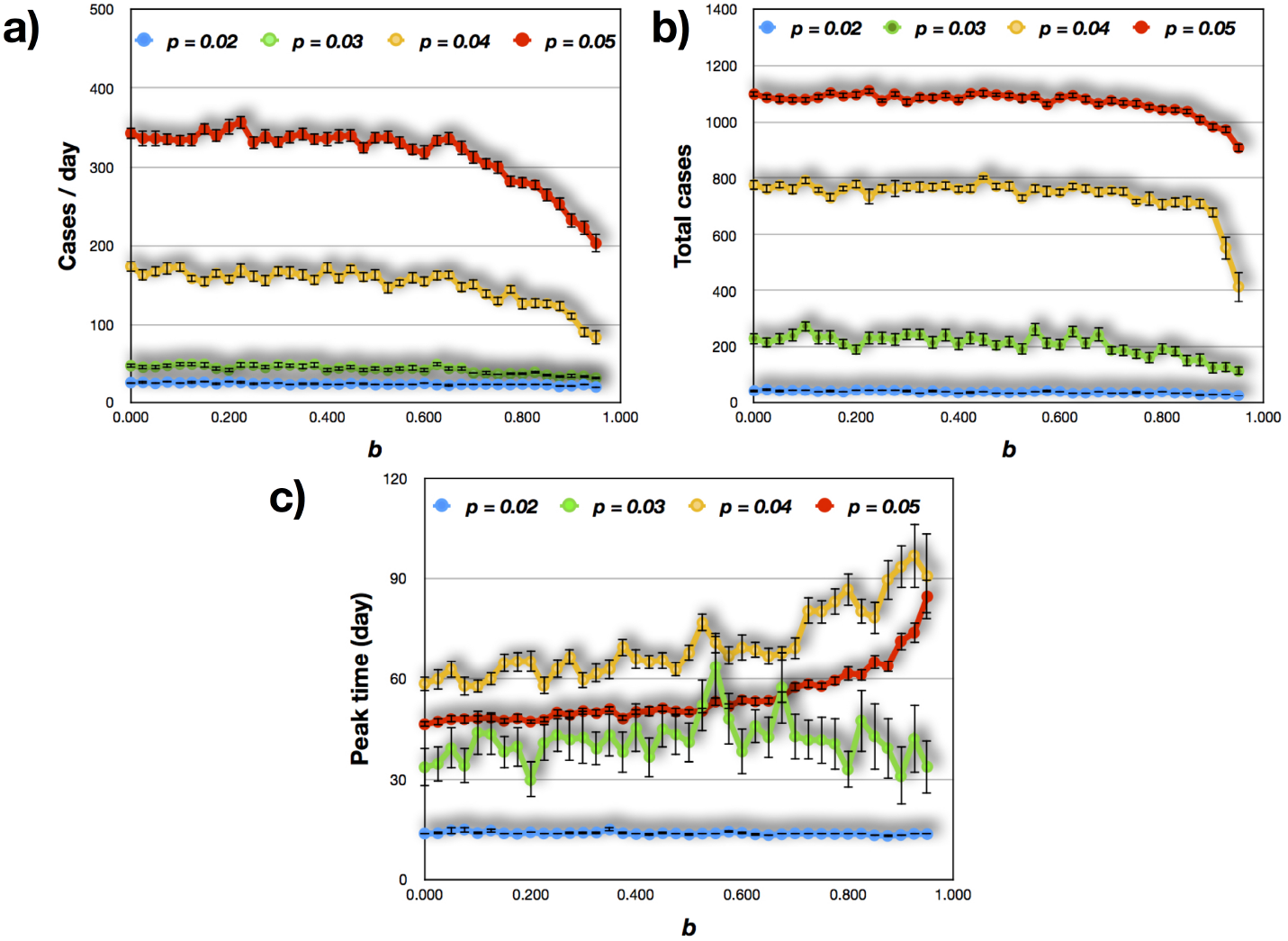
Cases per day, total cases and time in which the peak of infections occurs, depending on the magnitude of the barrier. In this case the barrier is located at *x* = 50. This implies that the area of zone 2 represents 75% of the total area.

**Figure 6.2:**
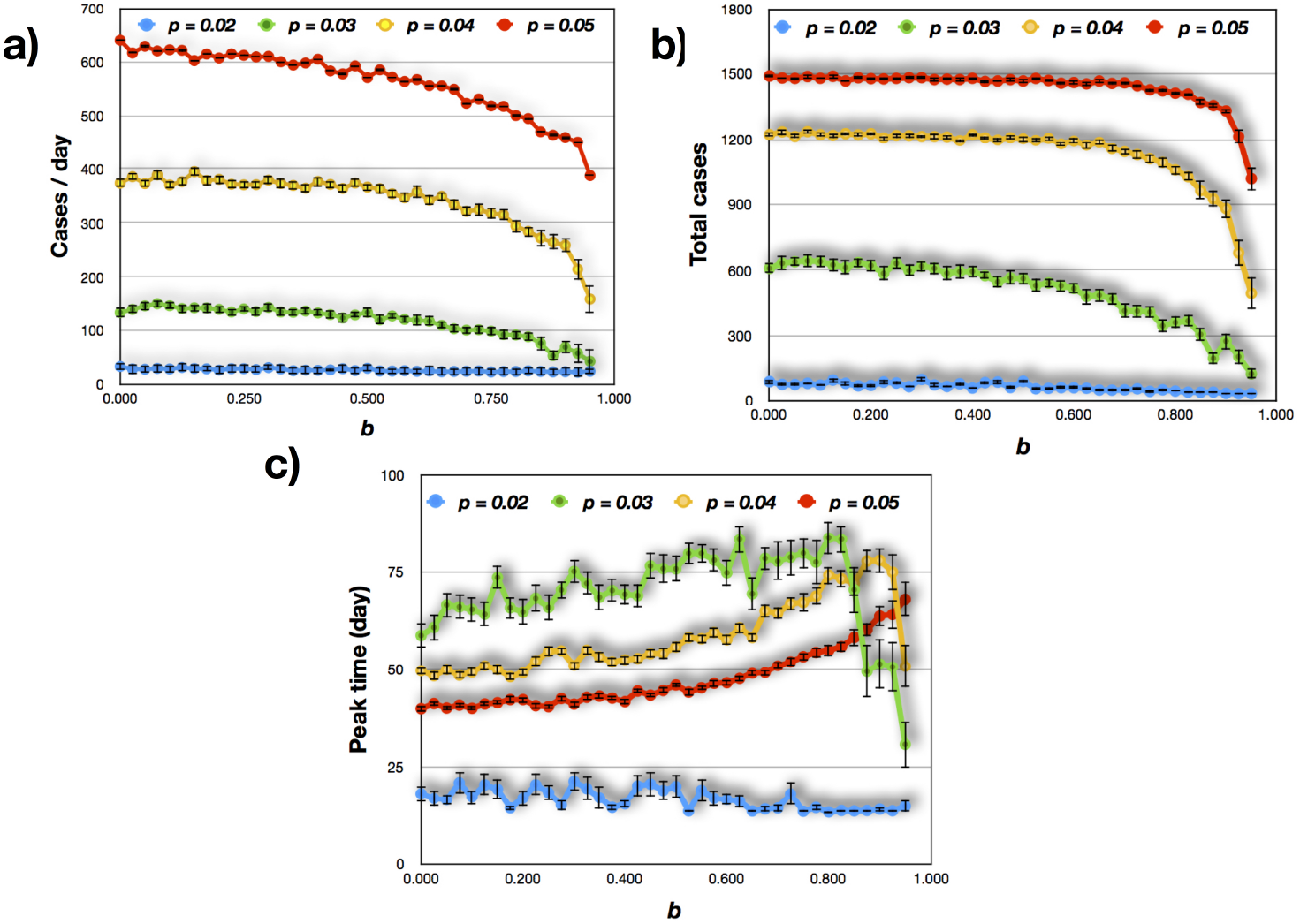
Cases per day, total cases and time in which the peak of infections occurs, depending on the magnitude of the barrier. In this case the barrier is located at *x* = 100. This implies that the area of zone 2 represents 50% of the total area.

**Figure 6.3:**
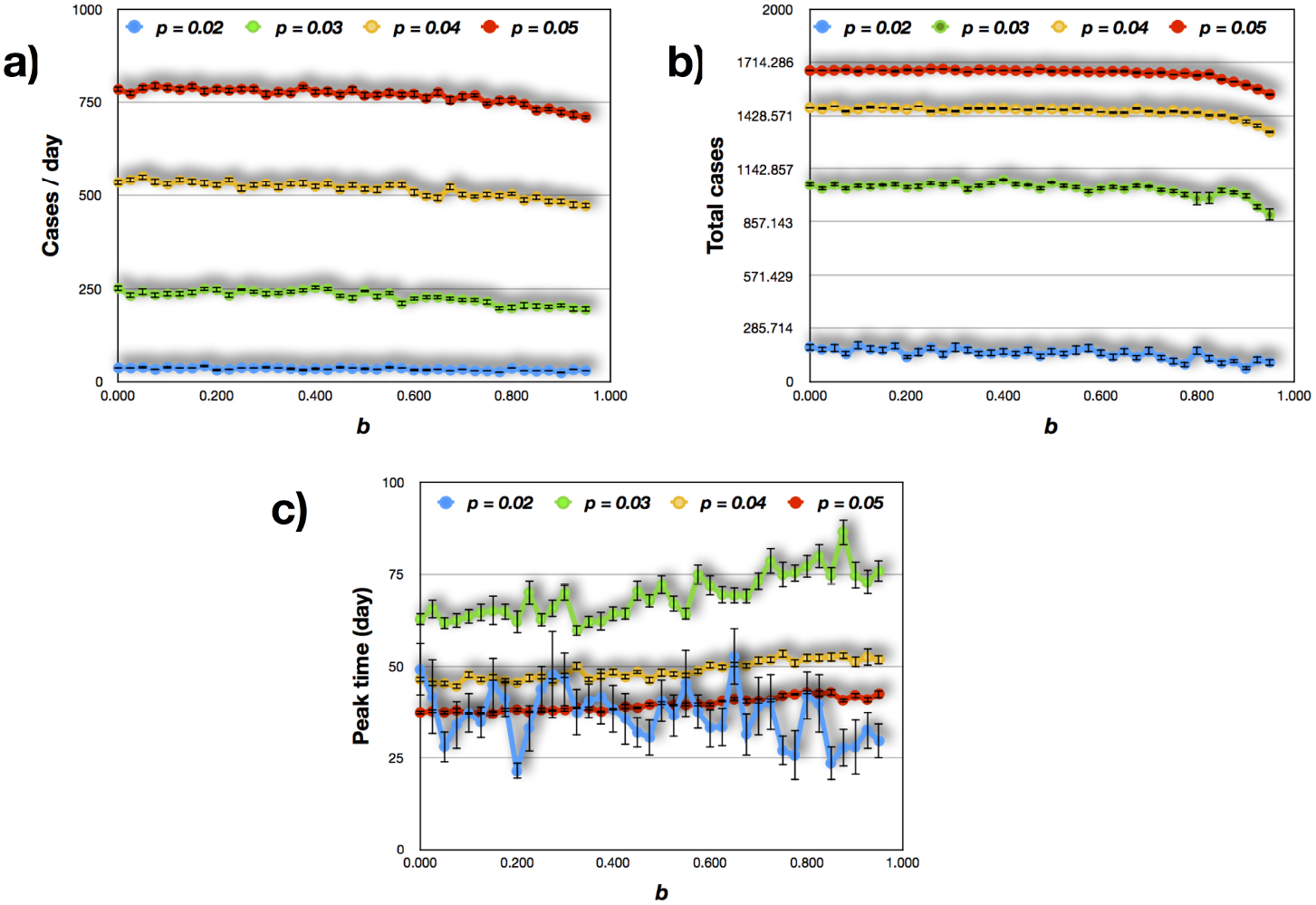
Cases per day, total cases and time in which the peak of infections occurs, depending on the magnitude of the barrier. In this case the barrier is located at *x* = 150. This implies that the area of zone 2 represents 25% of the total area.

This situation would indicate to us that under the partial quarantine regime, it would be more important to take into account the proportion of surface area and inhabitants of the areas that are divided and not in the sanitary barrier.

